# Microbiota composition in the lower respiratory tract is associated with severity in patients with acute respiratory distress by influenza

**DOI:** 10.1101/2021.12.07.21267419

**Authors:** Alejandra Hernández-Terán, Angel E. Vega-Sánchez, Fidencio Mejía-Nepomuceno, Ricardo Serna-Muñoz, Sebastián Rodríguez-Llamazares, Iván Salido-Guadarrama, Jose A. Romero-Espinoza, Cristobal Guadarrama-Pérez, JL Sandoval, Fernando Campos, Erika N. Mondragón-Rivero, Alejandra Ramírez-Venegas, Manuel Castillejos-López, Norma A. Téllez-Navarrete, Christopher E. Ormsby, Rogelio Pérez- Padilla, Joel A. Vázquez-Pérez

**Author notes:** **Corresponding author:** Joel Armando Vázquez-Pérez. These authors contributed equally (Alejandra Hernández-Terán, Angel E. Vega-Sánchez).

## Abstract

Several factors are associated with the severity of the respiratory disease caused by the influenza virus. Although viral factors are one of the most studied, in recent years the role of the microbiota and co-infections in severe and fatal outcomes has been recognized. However, most of the work has focused on the microbiota of the upper respiratory tract (URT), hindering potential insights from the lower respiratory tract (LRT) that may help to understand the role of the microbiota in Influenza disease. In this work, we characterized the microbiota of the LRT of patients with Influenza A using 16S rRNA sequencing. We tested if patients with different outcomes (deceased/recovered), use of antibiotics, and different days of symptoms onset differ in their microbial community composition. We found striking differences in the diversity and composition of the microbiota depending on the days of symptoms onset and with mortality of the studied patients. We detected a high abundance of opportunistic pathogens such as *Enterococcus, Granulicatella*, and *Staphylococcus* in patients either deceased or with antibiotic treatment. Also, we found that antibiotic treatment deeply perturbs the microbial communities in the LRT and affect the probability of survival in Influenza A patients. Altogether, the loss of microbial diversity could, in turn, generate a disequilibrium in the community, potentially compromising the immune response increasing viral infectivity, promoting the growth of potentially pathogenic bacteria that, together with altered biochemical parameters, can be leading to severe forms of the disease. Overall, the present study gives one of the first characterizations of the diversity and composition of microbial communities in the LRT of Influenza patients and its relationship with clinical variables and disease severity.

## Introduction

Several factors are associated with the severity of respiratory disease caused by influenza virus [1]. Overall, viral factors are one of the most studied and particularly, determinants of pathogenicity such as Hemagglutinin (HA), Non-structural protein 1 (NS1) and polymerases have been related with severity [2–5]. Other factors such as secondary bacterial infections [6] have been related with severity, as well as several other studies that have found bacterial co-infections associated with a more severe outcome or mortality during influenza epidemics in the last century [6–8].

Investigations into specific co-infecting pathogens that increase disease severity have shown that the most frequent bacteria are *Streptococcus pneumoniae, Haemophilus influenzae, Neisseria meningitidis and Staphylococcus aureus* [6]. In many cases, patients with co-infections present longer hospital stays and more severe disease [9]. Although these associations of co-infection and severity are well established, there is no conclusive evidence that all cases with severe or fatal outcomes have occurred in patients with co-infections, given that other clinical factors like obesity, hypertension, and other comorbidities have an important role in severity [9].

For the last ten years, modern technologies have allowed the study of whole microbial communities associated with hosts (microbiota) and is the subject of increasing interest [10,11]. In human hosts, most of the efforts have been focused on characterizing the microbiota in organs and tissues, such as the skin and the respiratory tract, but most have focused on digestive tissues and processes [12]. In the respiratory tract, there has been much progress describing and analyzing the diversity and composition of the microbiota in healthy and in pathological states. For instance, patients with chronic respiratory diseases such as COPD showed striking differences in the community composition of the respiratory tract compared with healthy controls [13,14]. In acute respiratory infections, and in particular influenza-like illness, there are several studies that characterize and analyze the respiratory microbiota [15–17]. Nonetheless, most of them have focused on the microbiota of the upper respiratory tract (URT) hindering potential insights from the lower respiratory tract (LRT) that may help in the understanding of the role of the respiratory microbiota in infectious diseases.

The composition of the microbiota has been recognized as an important factor in the homeostatic state of healthy individuals, it has been shown that an imbalance in the composition can cause or deepen the pathological conditions [18]. Factors such as age, gender, geographic region, diet, diseases, and antibiotic treatment can modify the composition and equilibrium of the microbiota, leading to a state known as dysbiosis [19–21]. Particularly, the respiratory microbiota plays a critical role in shaping the host’s immune response, which is essential for effective elimination of invading viruses [20,22–24]. Some studies have shown that healthy commensal microbiota help maintain a robust antiviral immunity, while dysbiosis increases viral infectivity due to the impaired capacity of the immune system to limit viral infection [25]. Recently, several reports point out that antibiotics can provoke dysbiosis in the gut and in the respiratory microbiota, causing different effects on homeostasis, microbial composition, and normal function against viruses [26].

In influenza disease, some reports have highlighted the importance of the microbiota and that antibiotics treatments could affect the microbiota response against viral infectious diseases [20,27]. For instance, it has been described that the microbiota present in mouse lung stromal cells induce an antiviral state driven by an interferon response. However, the antibiotic treatment has been found to affect this antiviral response in this model [20] and can even modify the influenza vaccine efficacy in humans [27].

In the present study, we aimed to characterize the microbiota of the LRT in patients with acute respiratory syndrome associated with influenza virus infection. Specifically, we tested if patients with different outcomes (deceased/recovered), use of antibiotics, and different days of symptoms onset differ in their microbial community composition and correlate with clinical data. We found striking differences in the diversity and composition of the microbiota depending on the days of symptoms onset and with mortality of the studied patients. We also found that the clinical use of antibiotics to reduce the severity of respiratory diseases was associated with an altered microbial community composition in the LRT which correlates with fatal outcomes.

## Methods

### Ethics statement

This study was reviewed and approved by the Science, Biosecurity and Bioethics Committee of the Instituto Nacional de Enfermedades Respiratorias (protocol number B0615) and the patients or their legal guardians gave their written consent.

### Sample collection

Endotracheal aspirates and bronchoalveolar lavages (BAL) were obtained from patients who were hospitalized at the Instituto Nacional de Enfermedades Respiratorias (INER) in Mexico City during the 2016-2017 and 2017-2018 winter seasons. Samples were taken from patients who were admitted to the emergency room with flu like symptoms (fever >38° C, and respiratory distress, with one or more of the subsequent symptoms: malaise, polypnea, cough or thoracic pain) and met the criteria for Acute Respiratory Distress Syndrome (ARDS) according to Berlin definition [28]. All samples were taken within the first 48 hours of hospital admission. Influenza A was detected using RT-PCR. Patients with more than 5 days of oseltamivir treatment and/or with chronic respiratory diseases (COPD, bronchiectasis) were excluded. A total of 30 patients met the inclusion criteria, and their demographic characteristics, severity scores, comorbidities, antibiotic/medication exposure, and clinical outcome were collected for all patients and shown in Table 1.

**Table 1.**
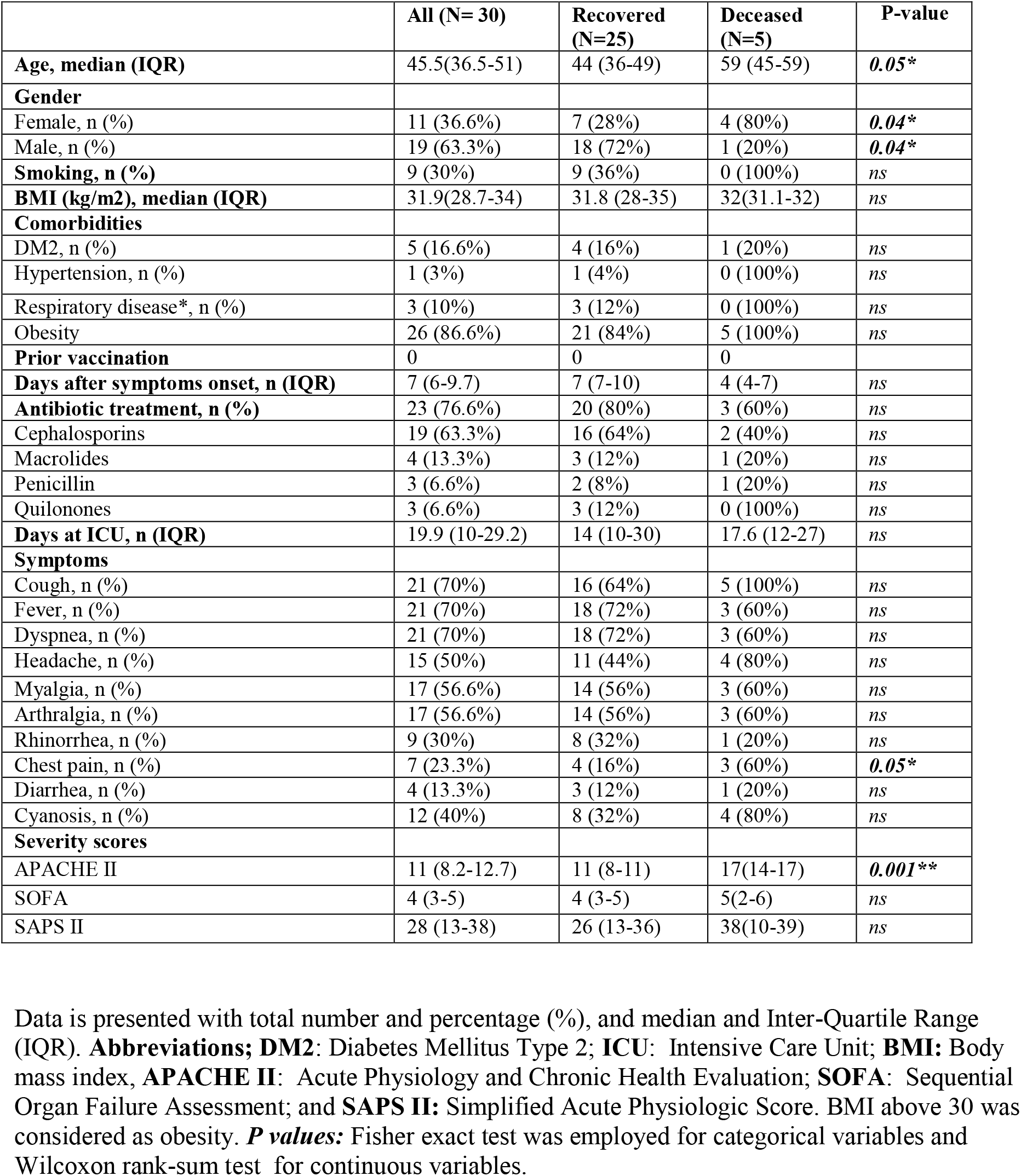
Characteristics and clinical data of the study subjects.

|  | All (N= 30) | Recovered (N=25) | Deceased (N=5) | P-value |
| --- | --- | --- | --- | --- |
| **Age, median (IQR)** | 45.5(36.5-51) | 44 (36-49) | 59 (45-59) | 0.05* |
| **Gender** |  |  |  |  |
| Female, n (%) | 11 (36.6%) | 7 (28%) | 4 (80%) | 0.04* |
| Male, n (%) | 19 (63.3%) | 18 (72%) | 1 (20%) | 0.04* |
| **Smoking, n (%)** | 9 (30%) | 9 (36%) | 0 (100%) | ns |
| **BMI (kg/m2), median (IQR)** | 31.9(28.7-34) | 31.8 (28-35) | 32(31.1-32) | ns |
| **Comorbidities** |  |  |  |  |
| DM2, n (%) | 5 (16.6%) | 4 (16%) | 1 (20%) | ns |
| Hypertension, n (%) | 1 (3%) | 1 (4%) | 0 (100%) | ns |
| Respiratory disease*, n (%) | 3 (10%) | 3 (12%) | 0 (100%) | ns |
| Obesity | 26 (86.6%) | 21 (84%) | 5 (100%) | ns |
| **Prior vaccination** | 0 | 0 | 0 |  |
| **Days after symptoms onset, n (IQR)** | 7 (6-9.7) | 7 (7-10) | 4 (4-7) | ns |
| **Antibiotic treatment, n (%)** | 23 (76.6%) | 20 (80%) | 3 (60%) | ns |
| Cephalosporins | 19 (63.3%) | 16 (64%) | 2 (40%) | ns |
| Macrolides | 4 (13.3%) | 3 (12%) | 1 (20%) | ns |
| Penicillin | 3 (6.6%) | 2 (8%) | 1 (20%) | ns |
| Quilonones | 3 (6.6%) | 3 (12%) | 0 (100%) | ns |
| **Days at ICU, n (IQR)** | 19.9 (10-29.2) | 14 (10-30) | 17.6 (12-27) | ns |
| **Symptoms** |  |  |  |  |
| Cough, n (%) | 21 (70%) | 16 (64%) | 5 (100%) | ns |
| Fever, n (%) | 21 (70%) | 18 (72%) | 3 (60%) | ns |
| Dyspnea, n (%) | 21 (70%) | 18 (72%) | 3 (60%) | ns |
| Headache, n (%) | 15 (50%) | 11 (44%) | 4 (80%) | ns |
| Myalgia, n (%) | 17 (56.6%) | 14 (56%) | 3 (60%) | ns |
| Arthralgia, n (%) | 17 (56.6%) | 14 (56%) | 3 (60%) | ns |
| Rhinorrhea, n (%) | 9 (30%) | 8 (32%) | 1 (20%) | ns |
| Chest pain, n (%) | 7 (23.3%) | 4 (16%) | 3 (60%) | 0.05* |
| Diarrhea, n (%) | 4 (13.3%) | 3 (12%) | 1 (20%) | ns |
| Cyanosis, n (%) | 12 (40%) | 8 (32%) | 4 (80%) | ns |
| **Severity scores** |  |  |  |  |
| APACHE II | 11 (8.2-12.7) | 11 (8-11) | 17(14-17) | 0.001* |
| SOFA | 4 (3-5) | 4 (3-5) | 5(2-6) | ns |
| SAPS II | 28 (13-38) | 26 (13-36) | 38(10-39) | ns |

Data is presented with total number and percentage (%), and median and Inter-Quartile Range (IQR). **Abbreviations;** **DM2:** Diabetes Mellitus Type 2; **ICU:** Intensive Care Unit; **BMI:** Body mass index, **APACHE II:** Acute Physiology and Chronic Health Evaluation; **SOFA:** Sequential Organ Failure Assessment; and **SAPS II:** Simplified Acute Physiologic Score. BMI above 30 was considered as obesity. **P values:** Fisher exact test was employed for categorical variables and Wilcoxon rank-sum test for continuous variables.

### DNA extraction and 16S rRNA sequencing

DNA was extracted using the QIAmp Cador Pathogen Mini Kit extraction (Qiagen N.V., Hilden, Germany) according to the manufacturer’s instructions. The V3-V4 16S rRNA region was amplified by PCR using the primers reported by Klindworth et al, (2013) [29] (F - 5’ CCTACGGGNGGCWGCAG 3’, R - 5’GACTACHVGGGTATCTAATCC 3’). Library preparation was carried out according to the Illumina 16S metagenomic sequencing protocol with minor modifications. Briefly, 16S amplicons were purified with the DNA clean & concentrator kit (Zymo Research, Irvine Cal., USA). Dual indices and Illumina sequencing adapters were attached in a second PCR step using Nextera XT Index Kit V2 (Illumina, San Diego Cal., USA). Finally, amplicons were purified, pooled in equimolar concentrations, and sequenced in a MiSeq Illumina instrument generating paired-end reads of 250bp.

### Sequence data processing

Illumina raw sequences were processed with QIIME2 (v.2020.8) [30]. Sequences denoising, quality filtering, and chimera detection were carried out with DADA2 [31]. After this process, an average of 34,516 per sample passed the cleaning and filter process. The Amplicon Sequence Variants (ASVs) were aligned with MAFFT [32] and used to construct a rooted tree for phylogenetic analysis with fasttree2 [33]. ASVs taxonomy was assigned with the *vsearch* classifier [34] using the Greengenes 13.8 database [35]. ASVs identified as mitochondria (N=13) and chloroplasts (N=375) were removed. Raw data were deposited in the NCBI Sequence Read Archive (SRA) (PRJNA770291).

### Compositional, diversity, and statistical analyses

Categorical variables were statistically compared using Fisher’s exact test and continuous variables using Wilcoxon rank-sum test. All statistical analyses were two-sided. All 16S analyses were performed using R v4.0.2 in RStudio v1.3.1 and the packages *ggplot2* (v3.3.3) [36], *vegan* (v2.5.7) [37], *microbiome* (v2.1.28) [38], *phyloseq* (v1.32.0) [39], and *randomcolorR* (v1.1.0.1) [40].

In order to compare the respiratory microbiota composition between different clinical features (e.g. previous antibiotic treatment, days of symptoms onset, and outcome) among Influenza A patients we performed the following analyses. For microbiota composition, we constructed stacked barplots in the *ggplot2* R package at phylum and genus level for all samples, and for the merged comparison groups in the *phyloseq* R package. Furthermore, we selected the top abundant genera for each group and compared its relative abundance using boxplots. Statistical differences in the abundance of such genera were calculated by a Wilcoxon rank-sum test in the *vegan* R package.

To compare alpha diversity among the analyzed groups we calculated diversity as the Shannon-Wiener index, and richness as the Chao1 index in with the *microbiome* R package and conducted a Wilcoxon rank-sum test with the *vegan* R package to detect statistical differences. For beta diversity we carried out a Principal Coordinates Analysis (PCoA) with weighted Unifrac distance at ASV level in the *phyloseq* R package. Potential differences in beta diversity were addressed with a Permutational Analysis of Variance (PERMANOVA) coupled with dispersion (PERMDISP) performed with the *vegan* R package.

Moreover, we also analyzed the clinical data associated with the patients included in this study. First, we performed a Canonical Analysis of Principal Coordinates (CAP) with weighted Unifrac distance in the *vegan* R package. We adjusted the model and plotted only non-redundant clinical variables. Also, to determine if the antibiotic treatment was correlated with the outcome, we constructed a Kaplan-Meier survival curve in SPSS Statistics (version 21) (Chicago, Illinois, USA) by using the days at the Intensive Care Unit (ICU) as time variable, the outcome (deceased or recovered) as a dependent variable, and the antibiotics treatment as exposure variable. Statistically significant difference was addressed with a Cox test. Finally, in order to detect if age and body mass index (BMI) affect alpha diversity in the respiratory microbiota, we performed Pearson correlations of those clinical variables and the diversity indexes (Shannon and Chao1).

## Results

### Study participants

A total of 30 Influenza A (H1N1) patients with flu-like disease and severe acute respiratory distress syndrome were analyzed. Demographic and health-related characteristics are described in Table 1. Briefly, the median age of our patient cohort was 45.5 (IQR: 36.5-51), 63.3% were male and 36.6% female. None of the patients received previous Influenza vaccination and most of them were either obese (86.6%) or present Diabetes Mellitus type 2 (DM2) (16.6%). 76.6% of the patients received antibiotic treatment prior to hospitalization. The median days of symptoms onset was 7 (IQR: 6-9.7). All patients were subject to Invasive Mechanical Ventilation (IMV). Regarding severity in terms of clinical indexes, the median value for the Acute Physiology and Chronic Health Evaluation II (APACHE II) was 11 (IQR: 8.2-12.7), for the Sequential Organ Failure Assessment index (SOFA) was 4 (IQR: 3-5), for the Simplified Acute Physiologic Score (SAPS II) was 28 (IQR: 13-38). Furthermore, deceased patients were significantly older (median 59, Fisher’s exact test, *p* = 0.05), mostly female (80%, Fisher’s exact test, *p* = 0.04), and with the highest APACHE II score (median 17, Fisher’s exact test, *p* = 0.001).

### Respiratory microbiota differs between deceased and recovered Influenza A patients

From the 30 respiratory samples taken from Influenza A patients we detected 11,083 Amplicon Sequence Variants (ASVs). At the phylum level (Fig 1A), Firmicutes dominated the composition of 83.3% of the samples, Bacteroidetes was found in the 80% of the samples, while other phyla such as Proteobacteria, Actinobacteria, Tenericutes, and Fusobacteria were detected in a small fraction of the samples and in less abundance (46.6%, 60%, 20%, and 16.6% respectively). In addition, we found that three samples were dominated entirely by Proteobacteria (INF430, INF437, and INF627) and one sample was dominated by Actinobacteria (INF857). Regarding the genus level (Fig 1B), it is important to highlight that all samples are composed by a small amount of different genera. *Veillonella, Streptococcus*, and *Prevotella* were found in almost all samples. While other genera such as *Erwinia, Haemophilus, Staphylococcus*, and *Oribacterium* were found in a big number of samples but in low abundance. In addition, there were some samples (almost all from deceased patients) dominated by known pathogens, such as *Acinetobacter* (INF437), *Pseudomonas* (INF627), *Rothia* (INF857), and *Granulicatella* (INF885) (Fig 1B).

**Figure 1.**
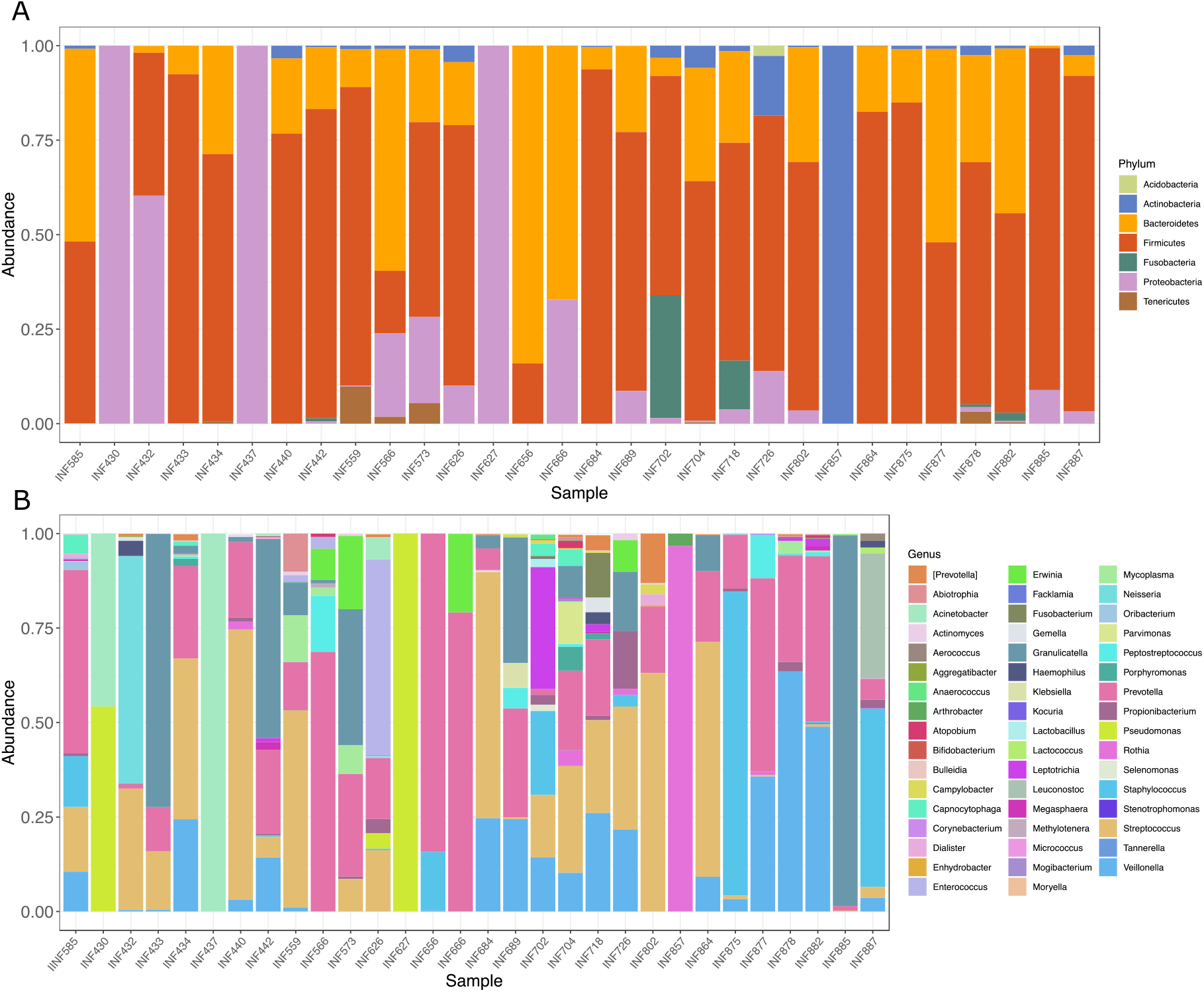
Composition of the respiratory microbiota across all patients at phylum and genus level. **A**. Stacked barplot depicting the relative abundance at phylum level for all samples. **B**. Stacked barplot depicting the relative abundance at genus level for all samples.

At comparing the microbial composition among deceased and recovered Influenza A patients we found important differences in particular microbial groups. For instance, we found that the whole respiratory microbial community of the deceased patients (Fig 2A) was composed by Proteobacteria, Bacteroidetes, and Firmicutes, clearly dominated by the last one. For recovered patients, we observed the presence of other phyla such as Tenericutes, Fusobacteria, and Actinobacteria. At the genus level (Fig 2B-C) we also appreciated differences in the dominance of specific microbes. For instance, in deceased patients the dominant genera were *Staphylococcus* and *Granulicatella*, with less abundance of other genera such as *Veillonella, Leuconostoc*, and *Erwinia*. In contrast, for the recovered patients we found a completely different arrangement in the community, being *Veillonella, Streptococcus*, and *Rothia* the dominant genera. Statistical differences in the abundance of these genera (e.g. *Veillonella, Streptococcus, Staphylococcus, Granulicatella, Prevotella*, and *Rothia*) between deceased and recovered patients are described in Figure 2C. It is worth to mention that although *Prevotella* appears in both groups, is significantly more abundant in recovered patients (Fig 2C). Moreover, we detected that recovered patients showed statistically more richness (Chao1 index, Wilcoxon rank-sum test, *p* = 0.01), and more diversity (Shannon index, Wilcoxon rank-sum test, *p* = 0.004) than deceased patients (Fig 2D). Finally, regarding beta diversity (Fig 2E), we also found differences in the spatial distribution in terms of weighted Unifrac distance of patients with different outcome (PERMDISP F= 7.71, *p* = 0.009).

**Figure 2.**
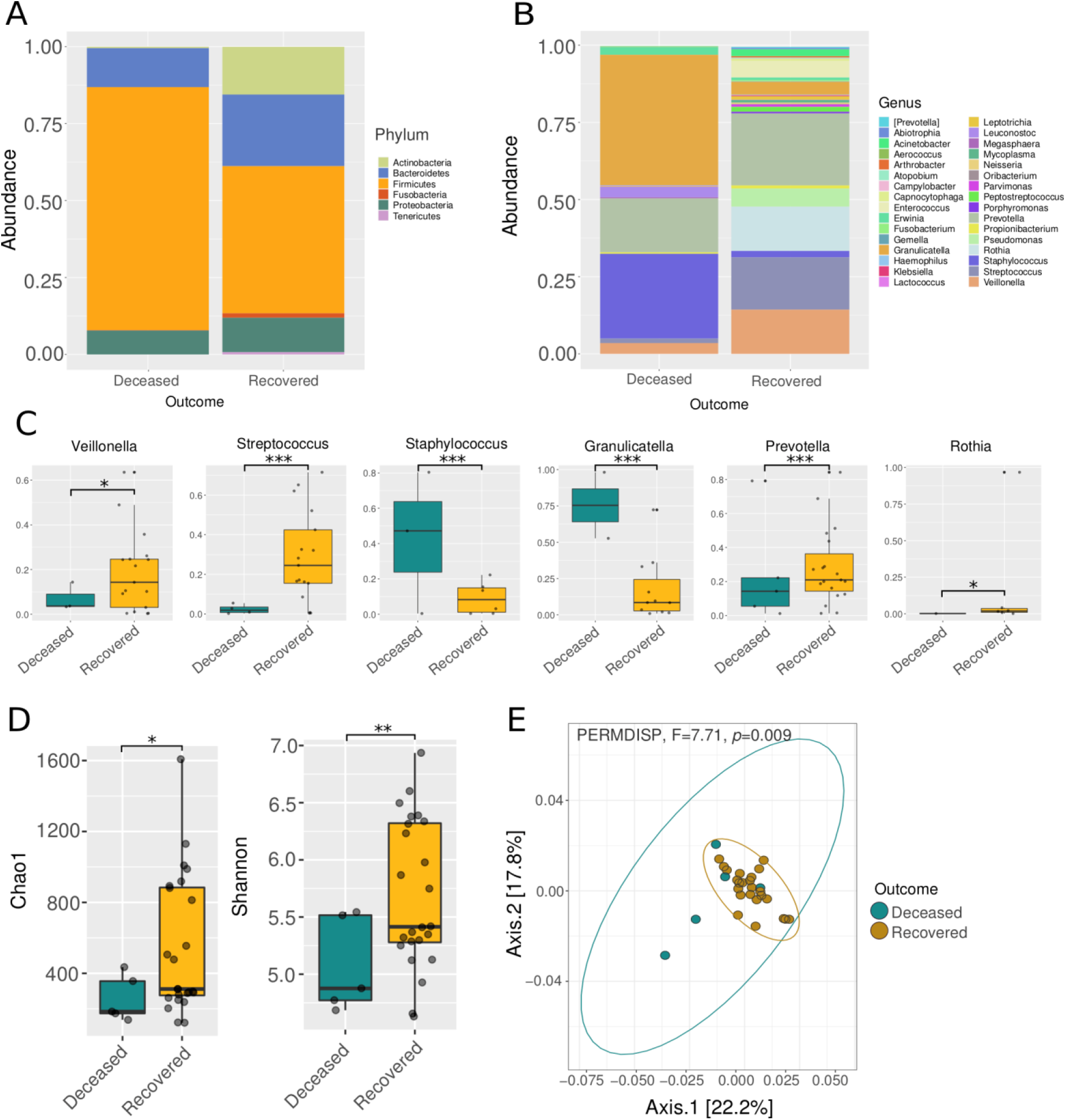
Diversity and composition of the respiratory microbiota in deceased and recovered patients with Influenza. **A**. Stacked barplot comparing the relative abundance of phyla between deceased and recovered patients. **B**. Stacked barplot comparing the relative abundance of genera between deceased and recovered patients. **C**. Boxplot of the relative abundance of the top abundant genera among the analyzed groups. **D**. Alpha diversity among the deceased and recovered patients. **E**. Principal Coordinates Analysis (PCoA) with weighted Unifrac distance and dispersion test (PERMDISP). Asterisks denote statistical significant differences given by a Wilcoxon rank-sum test (* *p* < 0.05, ** *p* < 0.001, *** *p* < 0.0001).

### Antibiotic treatment prior to hospitalization affect the diversity and composition of the respiratory microbiota in Influenza A patients

While analyzing the microbial communities between patients with and without previous antibiotic treatment we detected strong differences in the microbial composition. At the phylum level (Fig 3A) although Firmicutes and Bacteroidetes dominated the community in both groups (with (AB) and without (Non-AB) antibiotic treatment prior to hospitalization), only the AB patients showed high abundance of Actinobacteria. At the genus level (Fig 3B-C), the most abundant genera for both groups were *Veillonella, Streptococcus*, and *Prevotella*. While *Rothia, Prevotella, Granulicatella*, and *Enterococcus* were found significantly increased in patients with previous antibiotic treatment (AB, Fig 3C). Regarding alpha diversity (Fig 3D), we found significantly less richness (Chao1 index, Wilcoxon rank-sum test, *p* = 0.01) in the AB patients. In addition, we also found a significantly different arrangement (PERMDISP, F= 8.66, *p*= 0.006) of the microbial communities between AB and Non-AB patients according to the weighted UniFrac distance (Fig 3E).

**Figure 3.**
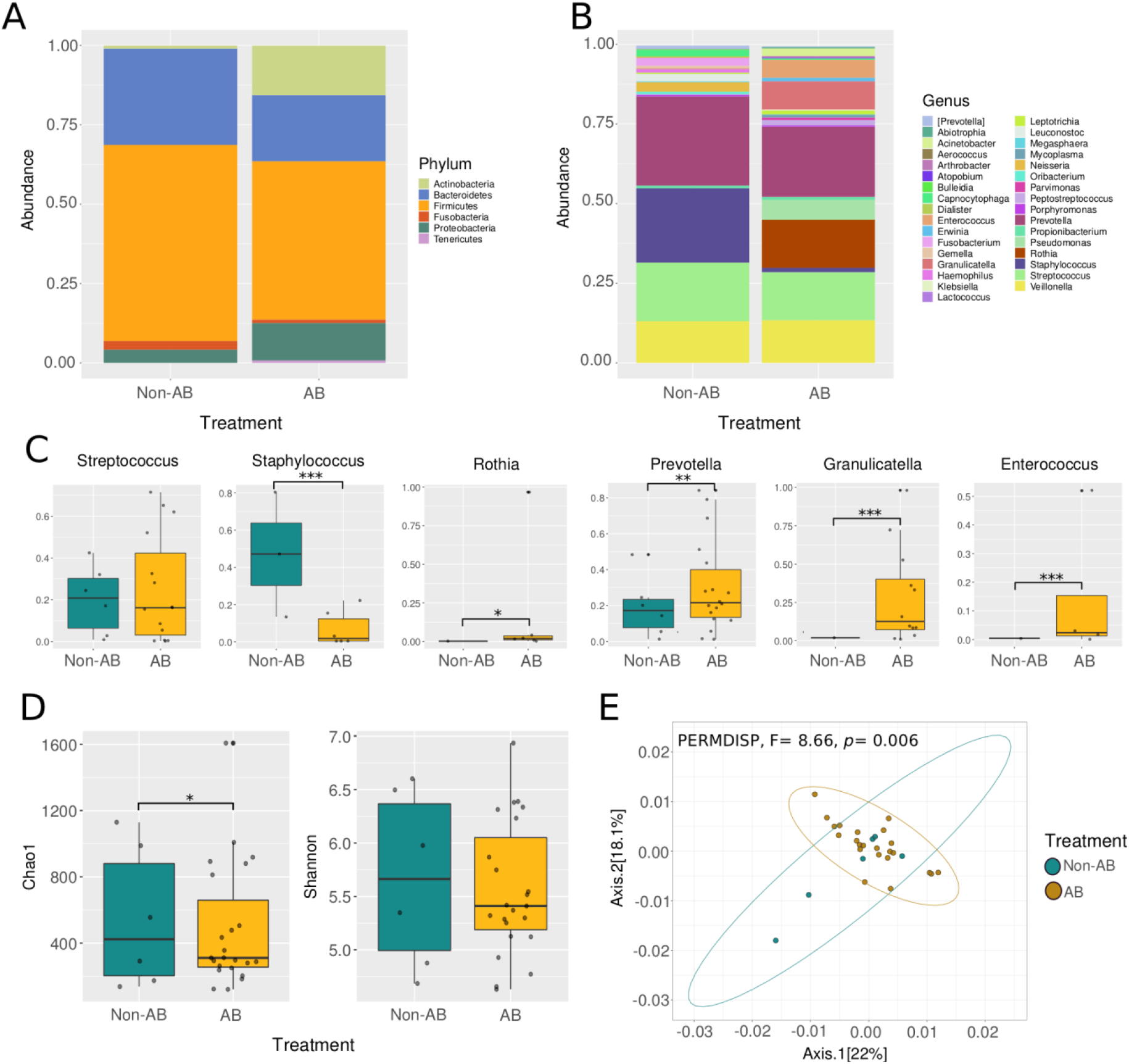
Diversity and composition of the respiratory microbiota among Influenza A patients with and without antibiotic treatment prior to hospitalization. **A**. Stacked barplot comparing the relative abundance of phyla between patients with (AB) and without (Non-AB) previous antibiotic treatment. **B**. Stacked barplot comparing the relative abundance of genera between patients with (AB) and without (Non-AB) antibiotic treatment. **C**. Boxplot of the relative abundance of the top abundant genera among the analyzed groups. **D**. Alpha diversity among AB and Non-AB patients. **E**. Principal Coordinates Analysis (PCoA) with weighted Unifrac distance and dispersion test (PERMDISP). Asterisks denote statistical significant differences given by a Wilcoxon rank sum test (* *p* < 0.05, ** *p* < 0.001, *** *p* < 0.0001).

### Patients with different days of symptoms onset differ in the microbial communities in lower the respiratory tract

When comparing patients with different days of symptoms onset we detected a distinct community in the respiratory tract between patients with early (0-8 days) and late (>9 days) symptoms onset (Fig 4). For the early symptoms patients (0-8 days), Actinobacteria, Bacteroidetes, and Firmicutes dominated the community at phylum level (Fig 4A). For the late symptoms patients (>9 days) we found the same phyla except for Actinobacteria, which appears absent. At the genus level (Fig 4B-C), we found *Veillonella, Streptococcus*, and *Enterococcus* significantly increased in the early symptoms patients (0-8 days), while *Granulicatella* and *Peptostreptococcus* were significantly increased in the late symptoms patients (>9 days). Regarding alpha diversity we do not find statistical differences among the compared groups (Fig 4D). Lastly, we found a slightly differentiation in beta diversity (Fig 4E) among the comparison groups (weighted UniFrac, PERMDISP, F= 6.5, *p*= 0.05).

**Figure 4.**
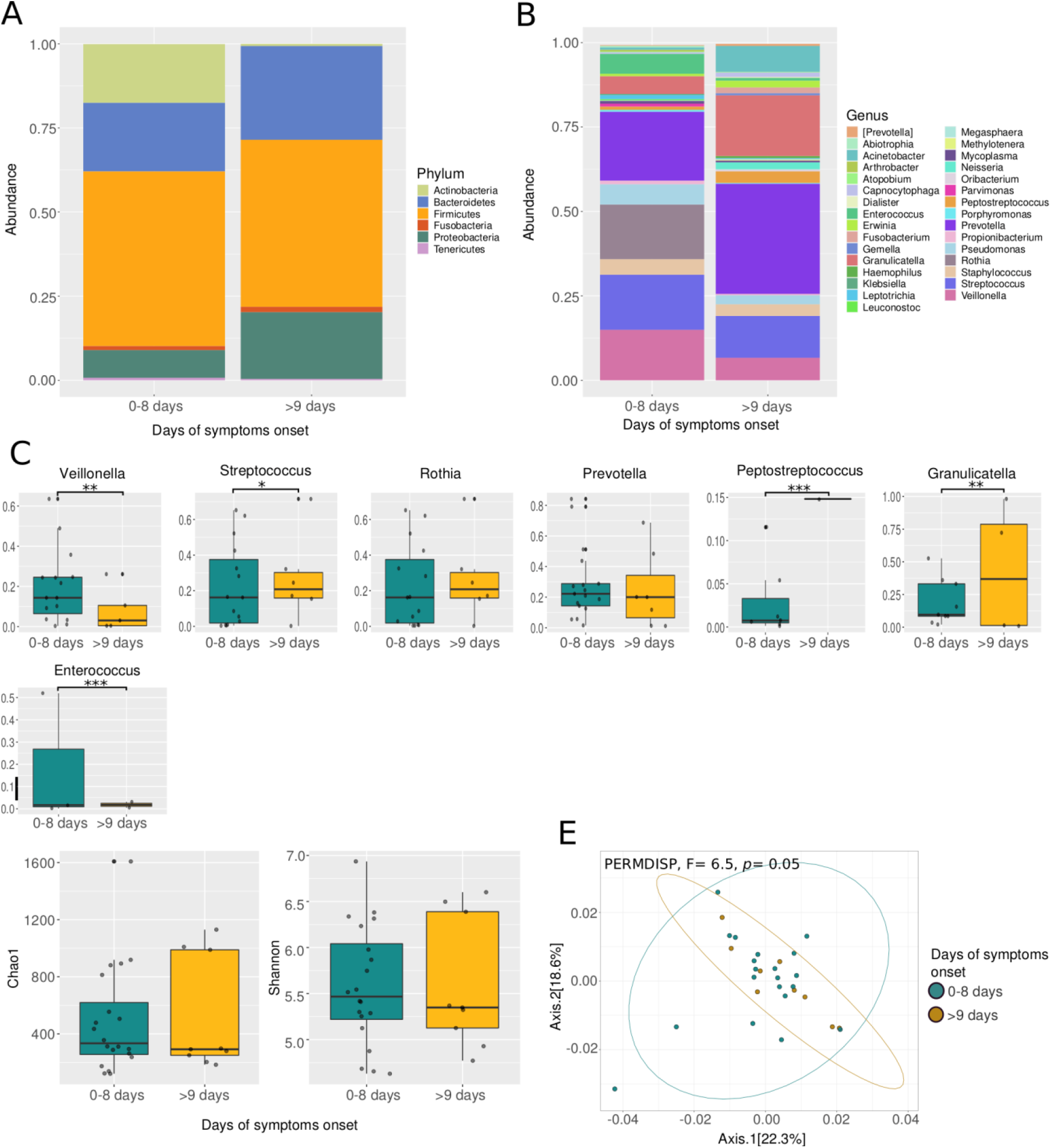
Diversity and composition of the respiratory microbiota among Influenza A patients with different days of symptoms onset. **A**. Stacked barplot comparing the relative abundance of phyla between patients early (0-8 days) and late symptoms onset (>9 days). **B**. Stacked barplot comparing the relative abundance of genera between patients early (0-8 days) and late symptoms (>9 days). **C**. Boxplot of the relative abundance of the top abundant genera among the analyzed groups. **D**. Alpha diversity among early (0-8 days) and late symptoms (>9 days) patients. **E**. Principal Coordinates Analysis (PCoA) with weighted Unifrac distance and dispersion test (PERMDISP). Asterisks denote statistical significant differences given by a paired Wilcoxon rank sum test (* *p* < 0.05, ** *p* < 0.001, *** *p* < 0.0001).

### Clinical features and respiratory microbiota in Influenza A patients

We found some clinical variables related with survival probability and microbial diversity and composition of the respiratory microbiota. For instance, after adjusting the CAP model, we found six clinical variables that together explain the 22% of the total variation (Fig 5A). We detected that all deceased patients were located in the upper half of the plot, positively correlating with lymphocytes, creatinine, and urea vectors (PERMANOVA, F= 1.9, *p* = 0.004). In contrast, although we found some recovered patients in the upper half plot, most of them were found in the lower half of the plot, correlating with total protein, and hematocrit vectors.

**Figure 5.**
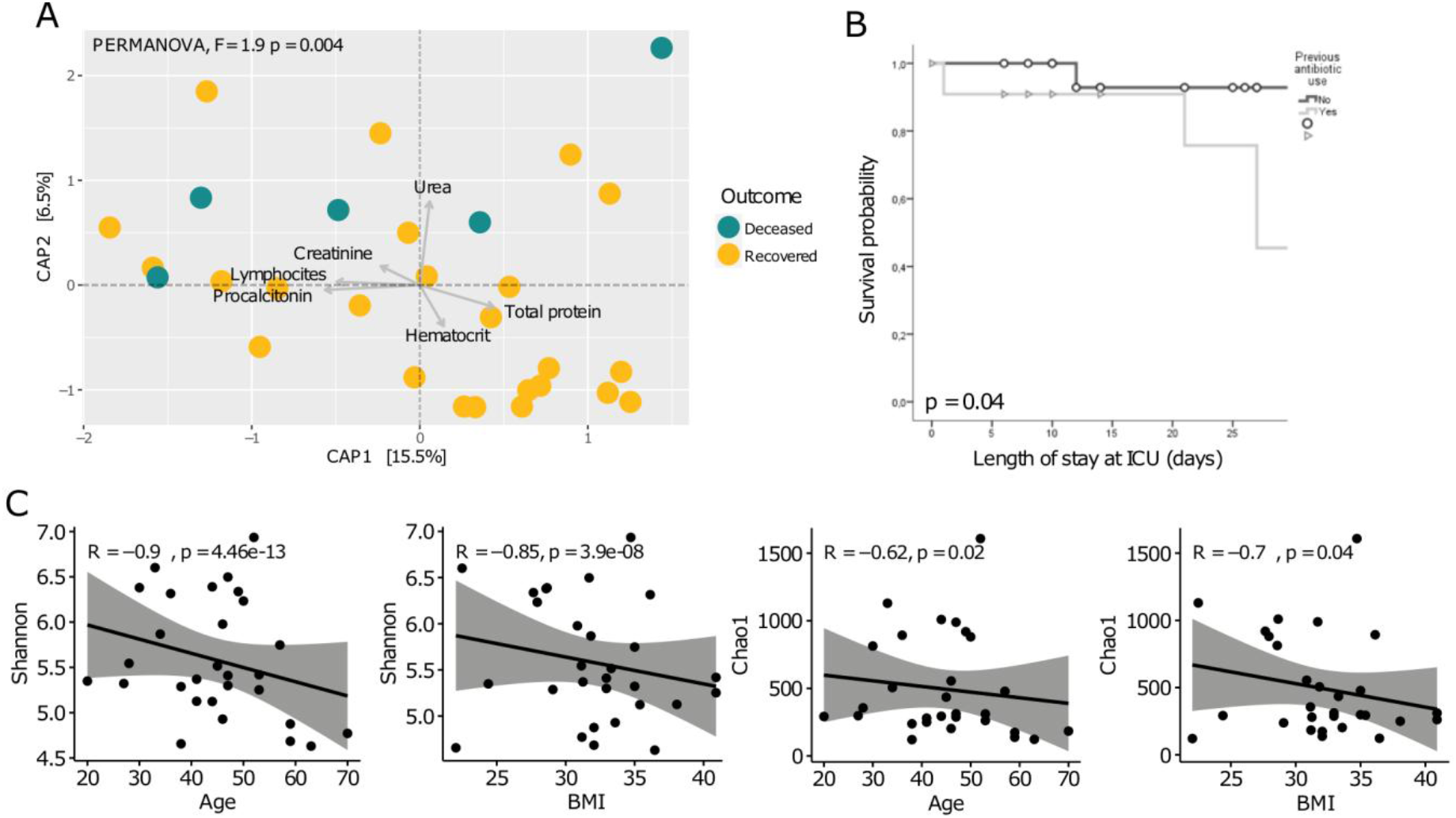
Clinical characteristics that correlate with microbial composition and survival probability. **A:** Canonical Analysis of Principal Coordinates (CAP) with weighted Unifrac distance and PERMANOVA test depicting clinical variables that contribute to explain variation. **B:** Kaplan-Meier survival curve with Cox test for antibiotic treatment and length of stay at ICU. **C:** Scatterplots illustrating Pearson correlations among diversity indexes (Shannon and Chao1) and age and Body Mass Index (BMI).

Furthermore, we found that patients with antibiotic treatment prior hospitalization exhibited less probability of survival than patients which were not treated with antibiotics (Fig 5B) (Kaplan-Meier curve, Cox test *p* = 0.04). Finally, we detected that, for all patients, both diversity (Shannon index) and richness (Chao1 index) were negatively correlated with age (R^2^ = -0.9, *p* = 4.4e-13 for Shannon index; R^2^ = -0.85, *p* = 3.9e-08 for Chao1 index) and BMI (R^2^ = -0.62, *p* = 0.02 for Shannon index; R^2^ = -0.7, *p* = 0.04 for Chao1 index) (Fig 5C).

## Discussion

The respiratory microbiota has proven to be related with disease courses in ARDS such as those caused by Influenza A virus [16,41,42]. In recent years, several studies have proposed that the microbiota in the LRT has an important role in lung morphology, function, and mucosal lymphoid tissue development, affecting the course of respiratory infectious diseases [43]. Nonetheless, due to the clinical limitations of sampling lungs (intubated patients) most of the work has been done in the URT (nasopharyngeal/oropharyngeal samples). Despite several studies trying to disentangle if microbial features on the URT could impact disease course (or the other way around) [8,16,41,42,44–46], only studies using mice models are available for the LRT [47].

Some studies have correlated the microbial composition in the respiratory tract with the outcome of patients with Influenza [7,20,45] and with other respiratory diseases [22,48]. On one hand, as it has been observed, patients with Influenza disease exhibit significantly low diversity compared to healthy controls [17,49]. In agreement with such previous studies of microbial composition in the respiratory tract, some subjects within our cohort exhibited a respiratory microbiota completely dominated by few genera, which is indicative of severe dysbiosis (Fig 1B). In many cases the dominating genera were microbial pathogens that have been previously found as co-infections with Influenza virus (e.g. *Pseudomonas, Acinetobacter*) [7,50].

It has been shown that the presence of potentially pathogenic bacteria could alter disease progression. At the phylum level, Firmicutes and Proteobacteria have been previously found to be enriched in patients with Influenza [25]. In accordance with Leung et al, (2013), we found Firmicutes and Proteobacteria notably increased in deceased patients, while in recovered patients, phyla such as Bacteroidetes and Actinobacteria were more abundant (Fig 2A). Regarding the genus level, in the microbiota associated to deceased patients we found *Granulicatella* and *Staphylococcus* to be enriched (Fig 2B-C), which have been associated to severe forms of Influenza in both children and adults [25,41,42]. Moreover, some species of the genus *Staphylococcus* are frequently found in co-infections with Influenza virus [6]. Thus, the commonly pathogenic bacteria frequently found associated with Influenza patients are particularly enriched in more severe forms of the disease and mortality within our cohort. Furthermore, we found that even between ill patients, the deceased patients showed significantly less richness (Chao1 Index) and diversity (Shannon-Wienner Index) than recovered patients (Fig 2D), meaning that a potential correlation between microbial diversity and outcome could exist.

It is known that antibiotic treatment affects microbial communities in hosts [19,27]. In mice pre-treated with antibiotics has been found an increased morbidity, mortality, and altered respiratory microbiota during H1N1 infection [20,23]. Particularly, it has been shown that antibiotics treatment can cause dysbiosis in the respiratory tract [26]. In this work, antibiotic treatment not only seems to cause alterations in the microbial communities as observed in other studies working with Influenza [27] and COVID-19 [51], but significantly affects the probability of survival (Fig 5B). In addition, patients that were prescribed antibiotics prior hospitalization showed, in accordance with previous reports [25,41,46,52], an increased abundance of potentially pathogenic bacteria such as *Granulicatella, Enterococcus, Streptococcus*, and *Rothia* (Fig. 3B-C) and significantly less richness (Chao1 Index, Fig. 3D) which could be in turn related with the fatal outcomes.

In order to disentangle the relevance of the respiratory microbiota on disease progression, we classified patients based on symptoms onset differences. We found important changes in the composition and abundance of microbial groups (Fig 4). Such alterations could be the result of the physiological changes that the host suffer along disease progression and that may impact the microbial communities. As the disease progresses, physiological and immunological processes take place and other clinical manifestations appear (hypoxemia, shock, renal failure among others) [53]. For instance, it has been reported that local immune response and inflammatory processes in advanced stages of the disease leads to acute increased mucus production [54] promoting anaerobic environmental niches in which bacteria such as *Granulicatella*, a known pathogen in oral cavities, can spread. Thus, viral replication and immunological mechanisms could be acting together causing the changes observed in patients with different days of symptoms onset, with potential consequences on the outcome.

Finally, we also test for associations between clinical features and microbial composition in the lower respiratory tract. For instance, the multivariate analysis (CAP) of clinical variables (Fig 5A) showed that some patients (in terms of Unifrac distance) were associated with laboratory variables related to severity in infectious diseases. In particular, some patients, including most of the deceased, were found positively correlated with urea, creatinine, and lymphocyte count. Deceased patients associated with elevated urea are of particular interest since such clinical parameter has been used as a marker for community-acquired pneumonia [55]. Moreover, both urea and creatinine are markers for acute kidney injury, which is a condition that has been associated with severe Influenza outcomes[56–58].

Furthermore, increased lymphocyte count has been associated with the exacerbated immune cellular responses that can occur in the acute phase of Influenza disease [59], a response that, to some degree, has been related to disruptions in the microbiota [23]. Also, we detected that deceased patients negatively correlated with total protein, which is generally found decreased in patients with advanced age, liver failure, and chronic inflammatory conditions (obesity, DM2, and hypertension) [60–62]. Additionally, we found that age and BMI negatively correlated with microbial diversity and richness (Fig 5C). In particular, those characteristics are risk factors for infectious diseases [63,64] by carrying a host systemic deterioration that may impact microbial communities. Specifically, both the decline of physiologic functions and immunosenescence that goes with age leads to a greater use of drugs (antibiotics among others), changes in lifestyle, and dietary intakes that have consequences in the microbial communities of both gut and respiratory tract [65,66]. Moreover, obesity entails multiple disruptions such as chronic inflammation and hormonal imbalance [67] which are associated with dysbiosis in the gut microbiome [68].

Taken together, the disequilibrium in the microbial communities of the LRT (low diversity and high abundance of potentially pathogenic bacteria) found in Influenza A patients could be altering some intrinsic functions of the microbiota. Firstly, the altered state as a result of antibiotics treatment may lead to an increased viral infection [25], by decreasing colonization resistance and affecting the containment of pathogens in the mucus [69]. As proven in other studies, generally Influenza A infection is accompanied with a disequilibrium of the URT. In murine models, it has been found that such alterations in the URT facilitate the entrance of bacterial groups (either commensal or pathogenic) to the LRT, thus promoting infection of lungs [50]. As a consequence, an alteration of the dynamic equilibrium in the lungs ecosystem could deeply affect antiviral response of the host [20,21], and even alter influenza vaccine efficacy [27].

## Conclusions

Overall, the present study gives one of the first characterizations of the diversity and composition of microbial communities in the LRT of Influenza patients and its relationship with clinical variables and disease severity. It is worth mentioning that given the nature of the analyzed samples, the main limitations of our study are the small number of subjects and the absence of healthy controls. In this work, we demonstrate that antibiotics and viral infection affect not only the URT but the microbial communities inhabiting the lungs. We found that antibiotic treatment deeply perturb the diversity and composition of the microbial communities in the LRT and the probability of survival in Influenza A patients. The loss of microbial diversity could, in turn, generate a disequilibrium in the community, potentially compromising the immune response increasing viral infectivity, promoting the growth of potentially pathogenic bacteria that, together with altered biochemical parameters, can be leading to severe forms of the disease. Finally, our findings support the idea that caution must be taken when antibiotic treatment is prescribed in patients with viral lung diseases.

## Data Availability

All data produced are available Raw data at the NCBI Sequence Read Archive (SRA)(PRJNA770291).

## Acknowledgments

We thank Eduardo Márquez García from Unidad de Biología Molecular, INER for technical assistance in Illumina sequencing. We also thank all physicians of the ICU for assistance with patient’s management.

## Author contributions

Conceptualization: AEVS, ARV, NATN, and JAVP. Collection of clinical data: SRL, CGP, JLS, and FC. Investigation: FMN, ENMR, and JAVP. Formal analysis: AHT and MC. Visualization and data curation: AHT. Interpretation of clinical data: AEVS, RSM, CEO, SRL, and RPP. Writing - Original draft preparation: AHT, AEVS, RSM, RPP, and JAVP. Writing - Review & Editing: all authors.

